# A proof-of-concept study: Investigating the impact of *COMT* genotype and proline on negative symptoms in Alzheimer’s Disease

**DOI:** 10.1101/2025.05.15.25327175

**Authors:** James D. Clelland, Hector W. Cure, Ashley M. Canizares, Julia H. Anderson, Bimala Rawal, Sabrina A. Wong, Nancy Kerner, Edward D. Huey, Lawrence S Honig, Devangere P. Devanand, Catherine L. Clelland

## Abstract

Previous studies have reported that levels of fasting plasma proline and catechol-O-methyltransferase (*COMT*) Val^158^Met genotype influence negative symptoms in patients with severe psychiatric illness and those at-risk for psychosis. Parallel negative neuropsychiatric symptoms are also a core feature of Alzheimer’s disease (AD). In this proof-of-concept cross-sectional study of dementia patients, we explored the relationship between *COMT* and proline on negative neuropsychiatric symptoms. The proline *x COMT* interaction significantly predicted symptoms as assessed via the negative items of the Positive and Negative Symptom Scale (n=50, interaction coefficient=0.025, p=0.031): Higher proline was beneficial for both Val/Val and Val/Met dementia patients, but detrimental to Met/Met patients. That high proline also has opposing effects on negative symptoms by *COMT* genotype in patients with dementia, further supports the development of therapeutics to specifically target the interaction pathway across neuropsychiatric disorders.

## Introduction

Negative neuropsychiatric symptoms, such as apathy (characterized by the loss of motivation to participate in activities, social withdrawal, and emotional indifference), blunted affect, and alogia, are a core feature of Alzheimer’s disease (AD), and constitute a behavioral dimension independent of depression and cognitive status^1-3^. Patients with mild cognitive impairment (MCI) also manifest negative neuropsychiatric symptoms^4^. Negative symptoms contribute substantially to the huge personal and economic costs of dementia: They are associated with functional deficits^5^, and a rapid course of both cognitive and functional decline^6,7^. Additionally, caregiver burden and distress has been significantly associated with the presence of negative neuropsychiatric symptoms in dementia patients^8,9^.

In previous work we demonstrated that fasting peripheral levels of the amino acid and likely CNS neuromodulator proline^10^, coupled with differential activity of the catechol-O-methyltransferase (COMT) enzyme, which metabolizes dopamine (as assessed via the *COMT* Val^158^Met functional polymorphism), interact to predict negative symptom outcomes in patients with^10^, or those at risk for^11^, severe psychiatric illnesses. For patients with one or two copies of the low-activity enzyme (*COMT* Met allele), who are likely to have high frontal cortical dopamine, high levels of plasma proline were associated with significantly greater negative symptom severity or less symptom improvement over time. Conversely, for those with high COMT activity (Val/Val), high plasma proline levels were associated with less severe negative symptoms, or greater symptom improvement^10,11^. Based upon these prior findings and evidence of proline elevation in AD^12-14^, we tested the hypothesis that *COMT* genotype and proline interact to modify negative symptom severity across neuropsychiatric disease diagnoses, specifically now investigating patients with dementia.

## Methods

We conducted a cross-sectional study of patients with a clinical diagnosis of probable AD or mild cognitive impairment with underlying AD biomarkers (MCI+). All procedures contributing to this work comply with the ethical standards of the relevant national and institutional committees on human experimentation and with the Helsinki Declaration of 1975, as revised in 2013. The protocol was approved by the Columbia University IRB (numbers AAAR8755, AAAS0071, and AAAV5130), and with the U.S. Army Medical Research and Development Command, Office of Human and Animal Research Oversight. Eligible subjects were identified from participants of Columbia University’s Alzheimer’s and Dementia Research Center (ADRC), Columbia’s Memory Disorders Clinic, and from patients under the care of the Columbia Doctors Neurology Aging and Dementia Practice. Written informed consent was obtained from all subjects.

Subjects were instructed to fast 8+ hours prior to their onsite single study visit. Negative symptoms were assessed via the Positive and Negative Symptom Scale (PANSS) and the Scale for the Assessment of Negative Symptoms in AD (SANS-AD)^5^. Positive PANSS symptoms were also evaluated. The Geriatric Depression Scale (GDS) and Mini-Mental State Examination (MMSE) were employed to assess depression and cognition, respectively. A fasting blood draw was obtained for assay of plasma proline and *COMT* genotyping, as described^10^.

Demographics, clinical characteristics, and medication status were compared across genotypes, and genotype distributions tested for Hardy-Weinberg equilibrium (HWE). The relationship between negative symptoms, demographics and clinical characteristics was also assessed. Testing the primary hypothesis of an interaction between *COMT* and proline on negative symptoms and following the statistical analysis design of our previous study^11^, we employed Least Absolute Shrinkage and Selection Operator (Lasso) regression for model prediction, using the double selection feature for testing *a priori* determined covariates (gender, depression, and positive symptoms). As a sensitivity analysis, a post-hoc stepwise selection procedure was employed, starting with the independent variables of proline, *COMT* and the *COMT x* proline interaction, plus Lasso retained covariates. Analysis was conducted in STATA/BE v18 (College Station, TX). For all tests, the level of significance was fixed at p<0.05, two-tailed. STROBE reporting guidelines were employed.

## Results

A total of 50 participants completed the study. Demographic and clinical characteristics are shown in Supplementary Table S1. The sample was in HWE for Val^158^Met (p>0.05), and subjects were well matched by genotypes. Regarding negative symptoms, there were no differences across genotypes for the negative items of the PANSS (sum of items N1, N2, G7, G8, G10)^15^, or the PANSS negative subscale (sum of items N1-N7). However, Val/Val subjects had significantly higher total SANS-AD scores (Table S1, p=0.044). As for our previous study of psychiatric patients^10^, we employed the negative items of the PANSS as the primary outcome variable, with secondary analyses of negative symptoms using the SANS-AD and the PANSS Negative Subscale. For all negative symptom assessments, significantly higher scores were observed in AD dementia as compared to MCI+ dementia (Supplementary Table S2), and therefore diagnosis was included as a covariate. Further, as expected subjects with AD had significantly lower MMSE scores compared to subjects with MCI+ dementia (Table S2, p=0.0006), and to avoid multicollinearity between the independent but related variables of diagnosis and cognition, diagnosis was chosen as the covariate for all subsequent models.

Via Lasso regression modelling, we observed a significant interaction between *COMT* and fasting plasma proline on negative symptoms (as assessed via the negative items of the PANSS), following adjustment for diagnosis and the retained variable of the PANSS positive subscale (interaction β-coefficient=0.025, p=0.031, Table 1). Higher proline was beneficial for both Val/Val and Val/Met dementia patients, but detrimental to Met/Met patients (Figure 1). In a post-hoc sensitivity analysis via a stepwise selection procedure, the interaction remained significant (Supplementary Materials Table S3), suggesting a robust finding. In secondary analyses, we tested for an interaction between *COMT* and proline on two additional and highly related measures of negative symptoms, the SANS-AD total score and the PANSS negative subscale; however, although supportive of our primary analysis, the interactions did not reach statistical significance (SANS-AD interaction β-coefficient=0.075, p=0.055, Table 1, Supplementary Figure S1: PANSS negative subscale interaction β-coefficient=0.030, p=0.093, additional model details not shown).

**Table 1.** Prediction of negative symptoms by the COMT x proline interaction, n=50

| Dependent / Retained Independent Variables | $\beta$ (95% CI) | se | z | p-value |
| --- | --- | --- | --- | --- |
| **Model 1: Negative Items (PANSS)<sup>a</sup> /** |  |  |  |  |
| Proline | -0.054 (-0.102, -0.007) | 0.024 | -2.24 | **0.025** |
| *COMT* | -6.510 (-12.06, -0.957) | 2.833 | -2.30 | **0.022** |
| *COMT* x proline | 0.025 (0.002, 0.047) | 0.011 | 2.16 | **0.031** |
| Diagnosis | -2.960 (-6.053, 0.133) | 1.578 | -1.88 | 0.061 |
| PANSS positive subscale | n/r |  |  |  |
| **Model 2: SANS-AD Total<sup>b</sup> /** |  |  |  |  |
| Proline | -0.165 (-0.318, -0.011) | 0.078 | -2.11 | **0.035** |
| *COMT* | -24.220 (-44.24, -4.202) | 10.21 | -2.37 | **0.018** |
| *COMT* x proline | 0.075 (-0.002, 0.153) | 0.039 | 1.92 | 0.055 |
| Diagnosis | -12.12 (-23.10, -1.135) | 5.602 | -2.16 | **0.031** |
| GDS total | 1.089 (-0.620, 2.798) | 0.872 | 1.25 | 0.212 |
| PANSS positive subscale | n/r |  |  |  |

<sup>a</sup>: Model 1, Wald $\chi^2(4) = 10.14$ , $p = 0.0382$ . Independent Variables: *COMT* (ordinal Val/Val, Val/Met, Met/Met), proline (continuous), interaction (*COMT* x proline), diagnosis (binary). Covariates: GDS total score (continuous), gender (binary), PANSS positive subscale (continuous).
<sup>b</sup>: Model 2, Wald $\chi^2(5) = 23.94$ , $p = 0.0002$ . Independent Variables: *COMT* (ordinal Val/Val, Val/Met, Met/Met), proline (continuous), interaction (*COMT* x proline), diagnosis (binary), GDS total score (continuous). Covariates: gender (binary), PANSS positive subscale (continuous).
n/r= point estimates of retained covariate(s) not reported in Lasso model.

**Figure 1.**
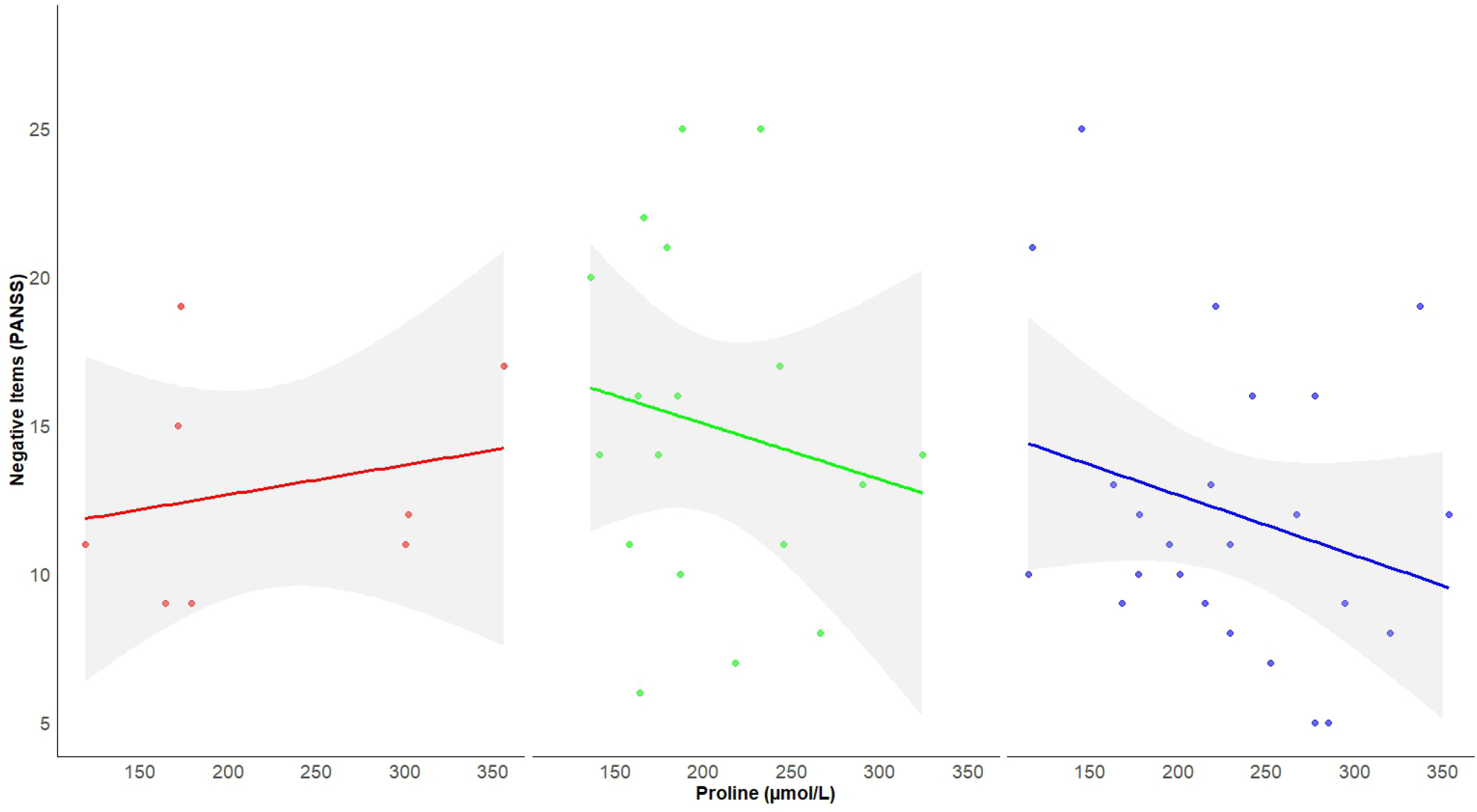
The Interaction between COMT Genotype and Proline on Negative Symptoms in Dementia patients. The graphs depict the interaction between COMT Val158Met genotype and proline on the negative items of the PANSS. The data is plotted for those with the Met/Met genotype (left panel, n=8), red), the Val/Val genotype (middle panel, n=18, green), and the Val/Met genotype (right panel, n=24, blue). Lines represent the predicted values from the simple regression models, with 95% confidence intervals. In those with the Met/Met genotype, high proline levels are associated with higher scores. Conversely there is a negative relationship in Val/Val and Val/Met patients, with high proline associated with fewer and less severe negative symptoms.

## Discussion

In this proof-of-concept study of patients with AD or MCI dementia with underlying AD biomarkers, we observed that levels of the CNS neuromodulator proline and *COMT* Val^158^Met genotype interact to predict negative neuropsychiatric symptoms. As for our studies of patients with severe psychiatric illness^10^ and those at-risk for psychosis^11^, higher proline was beneficial for Val/Val dementia patients, but detrimental to those with the Met/Met genotype. Elevated plasma proline (hyperprolinemia) has been associated with multiple neuropsychiatric phenotypes, including developmental delay and intellectual disability, autism spectrum disorder, and psychosis spectrum disorders^16,17^. There is also evidence of proline elevation in AD^12-14^. Further, in patients with 22q11 deletion syndrome, who routinely exhibit elevated proline due to hemizygous deletion of the *PRODH* gene on chromosome 22, negative neuropsychiatric symptoms show opposing severities by *COMT* allele^18-21^. Taken together we suggest a common mechanism across neuropsychiatric disorders: For Met/Met patients in the presence of high proline, a potential increase in dopamine transmission in the prefrontal cortex is exacerbated by low COMT activity, ultimately resulting in a frontal dopaminergic state above optimal levels. Conversely, in Val/Val patients, higher prefrontal COMT activity would likely reduce dopamine, limiting dopamine-receptor-mediated excitation^22,23^, and leading to potential prefrontal hypodopaminergia. In this instance, higher proline would be beneficial for Val/Val patients, via shifting prefrontal dopamine signaling to optimal levels. In this model, and as speculated^10,24^, negative symptoms would be induced in conditions of both hyper- and hypo-dopaminergia, reflecting the inverted U-shape curve proposed for cognitive dysfunction in psychosis^23^. We note that in this first study of dementia patients, high proline was in fact beneficial for Val/Met patients, which seems to be inconsistent with our prior findings on psychiatric patients^10,11^. It is interesting to speculate that reduced catecholamine levels in the aging cortex^25,26^, may dampen the impact of elevated proline, particularly for those patients with the Val/Met genotype, who have intermediate enzyme activity.

Although a limitation of this study is the small sample size, this first investigation of dementia patients builds positively from our previous studies. Furthermore, while the *COMT x* proline interaction did not reach statistical significance for all negative symptom assessments, we did observe a significant interaction using the negative items of the PANSS^15^, the one assessment that was used across all our studies^10,11^, with an interaction trending towards significance when measuring negative symptoms via the SANS-AD total score. However, replication of this work with a larger sample would be of benefit. Nevertheless, the finding that high proline has opposing effects on negative symptoms by *COMT* genotype in patients with dementia, further supports the development of therapeutics to specifically target the interaction pathway, across neuropsychiatric disorders.

## Acknowledgments

We would like to thank study coordinators and clinical staff of the Columbia University’s Taub Institute and ADRC, as well as Taub’s recruitment coordinators Bettina Idney and Lambrini Whitney. Most importantly, we would like to thank all the patients who participated in this study, as well as their caregivers.

## Author Contribution

Concept and design of the study-C. Clelland, J. Clelland. Acquisition, analysis, and interpretation of data-all authors. Statistical analysis-C Clelland. Drafting of the initial manuscript-C. Clelland. Critical revision and review of the final manuscript-all authors.

## Data Availability

The data that support the findings of this study are available from the corresponding author (C. Clelland), upon reasonable request, and approval by the Columbia IRB.

## References

1. Reichman WE, Coyne AC, Amirneni S, Molino B Jr, Egan S. (1996) Negative symptoms in Alzheimer’s disease. Am J Psychiatry. 153(3):424–6.

2. Galynker I, Ieronimo C, Miner C, Rosenblum J, Vilkas N, Rosenthal R. (1997) Methylphenidate treatment of negative symptoms in patients with dementia. J Neuropsychiatry Clin Neurosci. 9(2):231-9. Review.

3. de Jonghe JF, Goedhart AW, Ooms ME, Kat MG, Kalisvaart KJ, van Ewijk WM, Ribbe MW. (2003) Negative symptoms in Alzheimer’s disease: a confirmatory factor analysis. Int J Geriatr Psychiatry. 2003 Aug;18(8):748–53.

4. Hwang TJ, Masterman DL, Ortiz F, Fairbanks LA, Cummings JL. (2004) Mild cognitive impairment is associated with characteristic neuropsychiatric symptoms. Alzheimer Dis Assoc Disord. 18(1):17–21.

5. Benoit M, Andrieu S, Lechowski L, Gillette-Guyonnet S, Robert PH, Vellas B; REAL-FR group. (2008) Apathy and depression in Alzheimer’s disease are associated with functional deficit and psychotropic prescription. Int J Geriatr Psychiatry. 23(4):409–14.

6. Landes AM, Sperry SD, Strauss ME. (2005) Prevalence of apathy, dysphoria, and depression in relation to dementia severity in Alzheimer’s disease. J Neuropsychiatry Clin Neurosci. 17(3):342–9.

7. Lechowski L, Benoit M, Chassagne P, Vedel I, Tortrat D, Teillet L, Vellas B. (2009) Persistent apathy in Alzheimer’s disease as an independent factor of rapid functional decline: the REAL longitudinal cohort study. Int J Geriatr Psychiatry. 24(4):341–6. doi: 10.1002/gps.2125.

8. Fauth EB, Gibbons A. (2014) Which behavioral and psychological symptoms of dementia are the most problematic? Variability by prevalence, intensity, distress ratings, and associations with caregiver depressive symptoms. Int J Geriatr Psychiatry. 29(3):263–71. doi: 10.1002/gps.4002. Epub 2013 Jul 12.

9. Andrieu S, Coley N, Rolland Y, Cantet C, Arnaud C, Guyonnet S, Nourhashemi F, Grand A, Vellas B; PLASA group. (2015) Assessing Alzheimer’s disease patients’ quality of life: Discrepancies between patient and caregiver perspectives. Alzheimers Dement. pii: S1552-5260(15)02904-0. doi: 10.1016/j.jalz.2015.09.003.

10. Clelland CL, Drouet V, Rilett K, Smeed J, Nadrich RH, Rajparia A, Read LL, Clelland JD. (2016) Evidence that COMT Genotype and Proline Interact on Negative Symptom Outcomes in Schizophrenia and Bipolar Disorder. Translational Psychiatry, Sep 13;6(9):e891. doi: 10.1038/tp.2016.157.

11. Clelland JD, Hesson H, Ramiah K, Anderson JA, Thengampallil A, Girgis RR, Clelland CL. (2024) The Relationship Between COMT, Proline, and Negative Symptoms in Clinical High Risk and Recent Psychosis Onset. Transl Psychiatry 14, 409.

12. Pomara N, Singh R, Deptula D, Chou JC, Schwartz MB, LeWitt PA. (1992) Glutamate and other CSF amino acids in Alzheimer’s disease. Am J Psychiatry. 149(2):251–4.

13. Molina JA, Jiménez-Jiménez FJ, Vargas C, Gómez P, de Bustos F, Ortí-Pareja M, Tallón-Barranco A, Benito-León J, Arenas J, Enríquez-de-Salamanca R. (1998) Cerebrospinal fluid levels of non-neurotransmitter amino acids in patients with Alzheimer’s disease. J Neural Transm (Vienna); 105(2-3):279–86.

14. Trushina E, Dutta T, Persson XM, Mielke MM, Petersen RC. (2013) Identification of altered metabolic pathways in plasma and CSF in mild cognitive impairment and Alzheimer’s disease using metabolomics. PLoS One. May 20;8(5):e63644. doi: 10.1371/journal.pone.0063644.

15. Kane J, Honigfeld G, Singer J, Meltzer H. (1988) Clozapine for the treatment-resistant schizophrenic. A double-blind comparison with chlorpromazine. Arch Gen Psychiatry. Sep;45(9):789–96. doi: 10.1001/archpsyc.1988.01800330013001.

16. Vorstman JA, Turetsky BI, Sijmens-Morcus ME, et al. (2009) Proline affects brain function in 22q11DS children with the low activity COMT 158 allele. Neuropsychopharmacology 34(3):739–46.

17. Clelland CL, Read LL, Baraldi AN, Bart CP, Pappas CA, Panek LJ, Nadrich RH, and Clelland JD (2011) Evidence for association of hyperprolinemia with schizophrenia and a measure of clinical outcome. Schizophr Res. 131(1-3):139–45. Epub 2011 Jun 8.

18. Magnée MJ, Lamme VA, de Sain-van der Velden MG, Vorstman JA, Kemner C. (2011) Proline and COMT status affect visual connectivity in children with 22q11.2 deletion syndrome. PLoS One 6(10):e25882.

19. Zarchi O, Carmel M, Avni C, Attias J, Frisch A, Michaelovsky E, Patya M, Green T, Weinberger R, Weizman A, Gothelf D. (2013) Schizophrenia-like neurophysiological abnormalities in 22q11.2 deletion syndrome and their association to COMT and PRODH genotypes. J Psychiatr Res 2013; 47(11): 1623–9.

20. Hidding E, Swaab H, de Sonneville LM, van Engeland H, Vorstman JA. (2016) The role of COMT and plasma proline in the variable penetrance of autistic spectrum symptoms in 22q11.2 Deletion Syndrome. Clin Genet. 2016 Feb 26. doi: 10.1111/cge.12766. [Epub ahead of print]

21. Schneider M, Van der Linden M, Glaser B, et al. (2012) Preliminary structure and predictive value of attenuated negative symptoms in 22q11.2 deletion syndrome. Psychiatry Res. 196(2-3):277–84.

22. Bilder RM, Volavka J, Lachman HM, Grace AA. (2004) The catechol-O-methyltransferase polymorphism: relations to the tonic-phasic dopamine hypothesis and neuropsychiatric phenotypes. Neuropsychopharmacology 29(11):1943–61.

23. Tunbridge EM, Harrison PJ, Weinberger DR. (2006) Catechol-o-methyltransferase, cognition, and psychosis: Val158Met and beyond. Biol Psychiatry 60:141–151.

24. van Duin EDA, Ceccarini J, Booij J, et al. Lower [(18)F]fallypride binding to dopamine D(2/3) receptors in frontal brain areas in adults with 22q11.2 deletion syndrome: a positron emission tomography study. Psychol Med. 2020;50(5):799–807.

25. Wenk GL, Pierce DJ, Struble RG, Price DL, Cork LC. (1989) Age-related changes in multiple neurotransmitter systems in the monkey brain. Neurobiol Aging. 10(1):11–9. doi: 10.1016/s0197-4580(89)80005-3. PMID: 2569169 Review.

26. Zanto TP, Gazzaley A. (2019) Aging of the frontal lobe. Handb Clin Neurol. 163:369–389. doi: 10.1016/B978-0-12-804281-6.00020-3. PMID: 31590742 Review.

