## Supplementary Materials for "A proof-of-concept study: Investigating the impact of *COMT* genotype and proline on negative symptoms in Alzheimer’s Disease"

**Supplementary Methods:**

**Subjects.** Subjects aged 50 years or above, with a clinical diagnosis of probable Alzheimer’s disease (AD) or mild cognitive impairment with underlying AD biomarkers (MCI+), were eligible for recruitment. Subjects were recruited from Columbia University’s ADRC, Columbia’s Memory Disorders Clinic, or the Columbia Doctors Neurology Aging and Dementia Practice. Subjects were recruited between March 2019 and March 2025. Written informed consent was obtained from all subjects, prior to any procedures. 51 subjects were recruited; 50 completed the study with one subject dropped (as a clinical diagnosis could not be confirmed via medical records). The study was powered to detect an effect size >50% smaller than that found in Clelland et al, 2016^10^, with analysis stratified by gender. However, recruitment was impacted due to the COVID-19 pandemic and thus analyses performed using a non-stratified approach. Analysis of 48 subjects was determined to provide >85% power when α= 0.025, with three genotype groups plus covariates, suggesting that our study retained sufficient power to detect a significant interaction effect on negative symptoms across groups.

**Supplementary Results:**

**Supplementary Table S1. Demographic and clinical characteristics of the dementia subjects, n=50.**

| **Characteristic** | **Val/Val (n=18)** | **Val/Met (n=24)** | **Met/Met (n=8)** | **Prob**^a^ |
| --- | --- | --- | --- | --- |
| Gender, n (Female / Male) | 10 / 8 | 16 / 8 | 6 / 2 | 0.63 |
| Race, n |  |  |  | 0.11 |
| African American | 4 | 2 | 0 |  |
| Caucasian | 12 | 22 | 7 |  |
| Other / Not reported | 2 | 0 | 1 |  |
| Ethnicity, n (Hispanic or Latino, Y/N) | 7 / 11 | 8 / 16 | 2 / 6 | 0.85 |
| Age^b^ (years), mean ± SD | 72.2 ± 7.6 | 72.0 ± 8.0 | 66.1 ± 7.1 | 0.18 |
| Education^c^ (years), mean ± SD | 13.6 ± 4.4 | 15.4 ± 2.6 | 15.9 ± 3.3 | 0.19 |
| Testing Language (Engligh/Spanish) | 14 / 4 | 18 / 6 | 7 /1 | 0.90 |
| Diagnosis, n (AD/MCI+) | 15 / 3 | 19 / 5 | 7 / 1 | 1.00 |
| Fasting Plasma Proline^d^, mean ± SD | 203.3 ± 53.0 | 228.1 ± 65.2 | 220.5 ± 85.6 | 0.47 |
| **Symptoms** |  |  |  |  |
| Negative Items^e^ (PANSS), mean ± SD | 15.0 ± 5.8 | 12.1 ± 5.0 | 12.9 ± 3.7 | 0.20 |
| SANS-AD^f^ Total, mean ± SD | 37.1 ± 18.4 | 25.8 ± 18.0 | 19.8 ±15.2 | **0.04*** |
| PANSS Negative Subscale, mean ± SD | 21.5 ± 8.6 | 18.5 ± 8.1 | 19.0 ± 5.7 | 0.96 |
| PANSS Positive Subscale, mean ± SD | 11.3 ± 4.2 | 10.9 ± 4.1 | 11.3 ± 4.3 | 0.46 |
| GDS^g^ Total, mean ± SD | 3.6 ± 3.1 | 2.7 ± 3.0 | 2.5 ± 3.6 | 0.55 |
| MMSE^h^ Total, mean ± SD | 18.8 ± 6.6 | 19.8 ± 5.7 | 17.4 ± 4.2 | 0.60 |
| **Medications^i^** |  |  |  |  |
| Antidepressant, n (Y/N) | 6 / 11 | 7 / 17 | 3 / 5 | 0.85 |
| Antihypertensive, n (Y/N) | 9 / 8 | 8 / 16 | 4 / 4 | 0.40 |
| Antilipidemic, n (Y/N) | 9 / 8 | 9 / 15 | 1 / 7 | 0.16 |
| Cognitive Enhancer, n (Y/N) | 13 / 4 | 16 / 8 | 5 / 3 | 0.77 |
| Neuroleptic, n (Y/N) | 4 / 13 | 2 / 22 | 0 / 8 | 0.23 |
| Supplement, n (Y/N) | 10 / 7 | 14 / 10 | 4 / 4 | 0.93 |
| **a:** Significant p-value (<0.05) when comparing characteristics across three COMT genotype groups, calculated by one-way ANOVA, Kruskal-Wallis, or Fisher’s exact test, as appropriate. All tests were two-tailed.  **b**: n=49, as one subject did not report their age.  **c:** n=48, as two subjects did not report their years of education.  **d:** uMol/L  **e:** Items N1, N2, G7, G8, and G10 of the Positive and Negative Symptom Scale (PANSS)  **f:** Scale for the Assessment of Negative Symptoms in Alzheimer’s Disease.  **g:** Geriatric Depression Scale  **h:** Mini-Mental State Examination  **i:** n=49, as medication use for one subject was not available. Only medication groups prescribed to ≥10% of the sample (n=5) are reported. | | | | |

**Supplementary Table S2. Associations Between Demographic and Clinical Characteristics, with Negative Symptoms, n=50**

| **Characteristic** |  | **Negative Items (PANSS)** | **SANS-AD** |
| --- | --- | --- | --- |
| Gender | Male > Female | p=0.077 | p=0.095 |
| Ethnicity |  | p=0.379 | p=0.263 |
| Race |  | p=0.323 | p=0.247 |
| Age |  | p=0.899 | p=0.364 |
| Education |  | p=0.796 | p=0.365 |
| Testing Language |  | p=0.396 | p=0.310 |
| Diagnosis | AD > MCI+ | **p=0.026*** | **p=0.022*** |
| **Symptoms:** |  |  |  |
| PANSS Positive | Rho = 0.42 and 0.34 | **p=0.003*** | **p=0.016*** |
| GDS Total | Rho = 0.19 and 0.31 | p=0.183 | **p=0.028*** |
| MMSE Total | Rho = -0.55 and -0.39 | **p<0.001** | **p=0.005*** |
| **Medication:** |  |  |  |
| Antidepressant (Y/N) |  | p=0.271 | p=0.488 |
| Antihypertensive (Y/N) |  | p=0.606 | p=0.128 |
| Antilipidemic (Y/N) |  | p=0.675 | p=0.079 |
| Cognitive Enhancer (Y/N) |  | p=0.113 | p=0.167 |
| Neuroleptic (Y/N) | Y > N | p=0.067 | **p=0.015*** |
| Supplement (Y/N) |  | p=0.709 | p=0.835 |
| ***** Significant p-value when testing for an association with negative symptoms, calculated by T-test or one-way ANOVA for categorical variables, and Spearman correlations for continuous variables. When significant (p<0.05) or approaching a trend towards significance (p<0.1), the order or magnitude of the effect is documented. | | | |

**Supplementary Table 3. Post-Hoc Stepwise Section Modelling the *COMT x* proline interaction on Negative Symptoms, n=50**

| **Dependent / Retained Independent Variables** | | β (95% CI) | se | t | p-value |
| --- | --- | --- | --- | --- | --- |
| **Model 1:** Negative Items (PANSS) Total / | |  |  |  |  |
| Proline | | -0.057 (-0.110, -0.005) | 0.026 | -2.20 | 0.033 |
| *COMT* | | -7.024 (-12.96, -1.096) | 2.943 | -2.39 | 0.021 |
| *COMT* *x* proline | | 0.027 (0.003, 0.051) | 0.012 | 2.29 | 0.027 |
| PANSS Positive | | 0.526 (0.188, 0.863) | 0.168 | 3.13 | 0.003 |
|  | The stepwise section (criteria set to p<0.05) included the independent variables of proline, *COMT*, *COMT* *x* proline, and diagnosis, plus the Lasso variable of PANSS positive symptoms. Diagnosis was not retained in the model (Wald p>0.05). | | | | |

**Figure S1. The Interaction between *COMT* Genotype and Proline on Negative Symptoms in Dementia patients**. The graphs depict the interaction between *COMT* Val^158^Met genotype and proline on the SANS-AD total score. The data is plotted for those with the Met/Met genotype (left panel, n=8), red), the Val/Val genotype (middle panel, n=18, green), and the Val/Met genotype (right panel, n=24, blue). Lines represent the predicted values from the simple regression models, with 95% confidence intervals. In those with the Met/Met genotype, high proline levels are associated with higher scores. Conversely there is a negative relationship in Val/Val and Val/Met patients, with high proline associated with fewer and less severe negative symptoms.

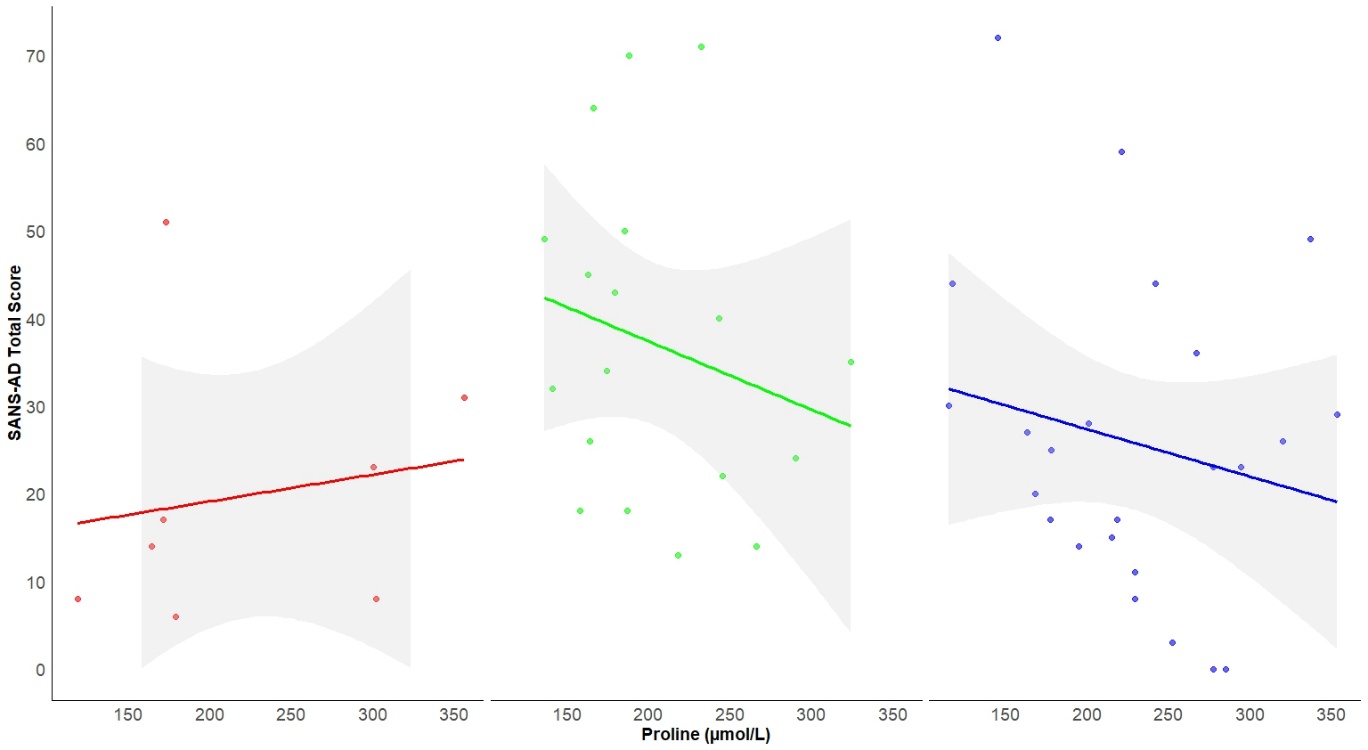
